# Post-COVID-19 syndrome and insulin resistance 20 months after a mild COVID-19

**DOI:** 10.1101/2023.04.17.23288637

**Authors:** Patricia Fierro, David Martín, Emilio Pariente, Ana B García-Garrido, Héctor Basterrechea, Benedetta Petitta, Camila Bianconi, Sara Herrán, Andrea Berrueta, Ascensión Jorrín, Alicia Gómez, Raquel Casado, Alfredo Cuadrado, Carmen Ramos, José L Hernández

**Author notes:** **CORRESPONDING AUTHOR** Emilio Pariente, MD, PhD, “Camargo Interior” Primary Care Center Associate Professor, University of Cantabria, Avda Bilbao, s/n. 39600-Muriedas, Cantabria, Spain. **FUNDING** The study has been carried out without any funding. **AVAILABILITY OF DATA** The datasets generated during and/or analysed during the current study are available from the corresponding author on reasonable request. **ETHICAL CONSIDERATIONS** The tenets of the Declaration of Helsinki in research on human subjects were followed. All patients who met the inclusion criteria were informed of the purpose of the study and were invited to participate. All of them expressed their verbal consent and there was no refusal to participate. The study was approved by the Clinical Research Ethics Committee of Cantabria (code 2021.102).

## Abstract

**Objective:** SARS-CoV-2 infection is associated with impaired glucose metabolism. Although the mechanisms are not fully understood, insulin resistance (IR) appears to be a central factor. Patients who had a severe acute phase, but even asymptomatic or with mild COVID-19, have an increased risk of T2DM. After the acute phase, post-COVID-19 syndrome (PCS) also seems to be related to this metabolic disturbance, but there is a paucity of studies. This study aims to evaluate a possible relationship between PCS and IR after mild COVID-19 and, if confirmed, whether there are differences by sex.

**Subjects and methods:** Retrospective observational cohort study including subjects who had mild COVID-19 between April and September 2020 in a community setting. None had been vaccinated against SARS-CoV-2 at inclusion, and previous T2DM and liver disease were exclusion criteria. Patients who met NICE criteria were classified as PCS+. Epidemiological and laboratory data were analysed. Three assessments were performed: 1E (pre-COVID-19, considered baseline and reference for comparisons), 2E (approximately 3 months after the acute phase), and 3E (approximately 20 months after the acute phase).

A triglyceride-to-glucose (TyG) index ≥8.74 was considered IR. Albumin-to-globulin ratio (AGR) and lactate dehydrogenase (LDH) were assessed as inflammatory markers. Bivariate analyses were performed, using nonparametric and repeated measures tests.

A subsample without metabolic disorder or CVD (age<median, BMI<25 kg/m2, elevated AGR, TyG index=7.80 [0.5]) was generated to reasonably rule out prior baseline IR that could bias the results. The relationships between PCS and TyG in 3E (TyG3) were modeled in 8 multiple regressions, stratifying by sex and BMI combinations.

**Results:** A total of 112 subjects (median [IQR] of age= 44 [20] years; 65 women) were analysed. Up to 14.3% was obese and 17% was hypertensive. Significant increases between 1E and 3E were registered regarding (i) basal glycemia (BG), 87 [14] mg/dL vs. 89 [14]; p=0.014, (ii) TyG index (8.25 [0.8] vs. 8.32 [0.7]; p=0.002), and (iii) LDH in 3^rd^ tertile (16.1% vs 32.1%; p=0.007). A total of 8 previously normoglycemic subjects, showed BG2 or BG3 >126 mg/dL.

The subgroups with IR highest prevalence at 3E were those of BMI ≥25 kg/m^2^ and PCS+. The subgroup without CVD presented a significant increase in the TyG index (TyG1=7.80 [0.1] vs. TyG3= 8.28 [0.1]; p=0.017). LDH1 was significantly correlated with TyG3 in both sexes (rho=0.214 in women, rho=0.298 in men); in contrast, LDH2 and LDH3 did not present such an association.

In multivariable analysis, PCS has shown to be an independent and predictive variable of TyG index in women with BMI<25 kg/m^2^, after adjustment for age, hypertension, BMI, Charlson comorbidity index, AGR1, AGR2, LDH1, number of symptoms of acute COVID-19, and number of days of the acute episode (β=0.350; p=0.039).

**Conclusions:** PCS has played a secondary role in predicting IR, showing a modest effect compared to BMI or prior hypertension. A significant increase in IR has been noted 20 months after mild COVID-19, both in cases of previous baseline IR and in those without previous IR. Basal serum LDH has shown to be predictive of current TyG, regardless of elevated LDH after SARS-CoV-2 infection. There were profound differences between women and men, confirming the need for a sex-stratified analysis when addressing the relation between PCS and glycemic alterations.

## INTRODUCTION

SARS-CoV-2 infection and type 2 diabetes mellitus (T2DM) have a bidirectional relationship [1]. From the early stages of the pandemic, T2DM and a hyperglycemic state were observed to increase the risk of a severe disease course [2]. Patients with T2DM have an immune system imbalance and a higher level of inflammatory biomarkers compared to patients without diabetes. An important role in this mechanism is attributed to Th17 and Treg cells, along with the secretion of inflammatory factors [3]. In addition to DM-associated low-grade inflammation affecting peripheral insulin sensitivity (IS), other mechanisms driven by hyperglycemia may be involved, such as elevated synthesis of advanced glycation end products (AGEs) [4] or direct stimulation of viral proliferation [1].

On the other hand, it has been observed that some non-diabetic patients with COVID-19 develop diabetes or have acute complications of pre-existing diabetes, including diabetic ketoacidosis. Pancreatic islets have a high expression of ACE2 receptors, and the high affinity of SARS-CoV-2 for them may cause direct β-cell injury [1] or indirectly, increased angiotensin II production, with fibrotic and inflammatory effects, and increased insulin resistance (IR) [3,5]. IR plays a central role in the pathogenesis of T2DM. Coexistence of other underlying processes, such as stress hyperglycemia or corticosteroid-induced hyperglycemia, is also possible. Hypovitaminosis D may in turn cause IR, which may be aggravated by high viral titres that stimulate the release of cytokines/chemokines, most of them with a negative effect on systemic IS [4].

Acute phase severity is associated with a significant increase in the incidence of T2DM, as demonstrated by several cohort studies [6]. However, even asymptomatic or mild COVID-19 patients who did not require hospitalization also have a significantly increased risk of T2DM [6].

After the acute COVID-19, many patients go on to develop lasting symptoms that fluctuate over time and can have disabling consequences. The most common long-term symptoms are chronic fatigue, persistent dyspnea and shortness of breath, general neurological decay and cognitive dysfunction. This condition, called post-COVID-19 syndrome (PCS), significantly affects patients’ quality of life, causes economic and productivity losses and increases the burden of care [7,8].

There is some evidence that PCS is a risk factor for DM [8]. Accumulating results point to several mechanisms that could be implicated: destruction of exocrine and endocrine cells due to SARS-CoV-2 infection, trandifferentiation of pancreatic beta cells through the activation of eIF2 signaling pathway, and induced autoimmunity and low-grade inflammation [4]. Of them, IR seems to be a pivotal factor, adscribed to low-grade inflammation [8]. Nevertheless, the pathophysiological and molecular mechanisms connecting PCS and T2DM are far from completely understood [4] and there is a paucity of studies addressing this relation [8]. Additionally, a previous study by our working group showed that subjects with PCS, along with slight but significant elevations of inflammatory markers, had marked sex differences in the inflammatory response [9].

Based on these considerations, this study aims to ascertain a possible relation between PCS and IR in outpatients after mild COVID-19 and, if confirmed, whether there are differences by sex.

## METHODS

### Design

Retrospective observational cohort study, which included subjects with a mild episode of COVID-19 in a community setting. The general characteristics are shown in **Figure 1**. Three evaluations, named 1E, 2E and 3E, were performed. ‘1E’ was an assessment prior to the acute episode of COVID-19, in which laboratory parameters were analysed in the context of a periodic health examination. It has provided the baseline analytical data of the participants and has been the reference for comparisons. ‘2E’ was a clinical and laboratory evaluation, whose details have been published [9], and has shown the clinical-metabolic situation after acute COVID-19. Finally, ‘3E’ has provided laboratory data 20 months after the acute episode, and is assumed to be the present.

**Figure 1:**
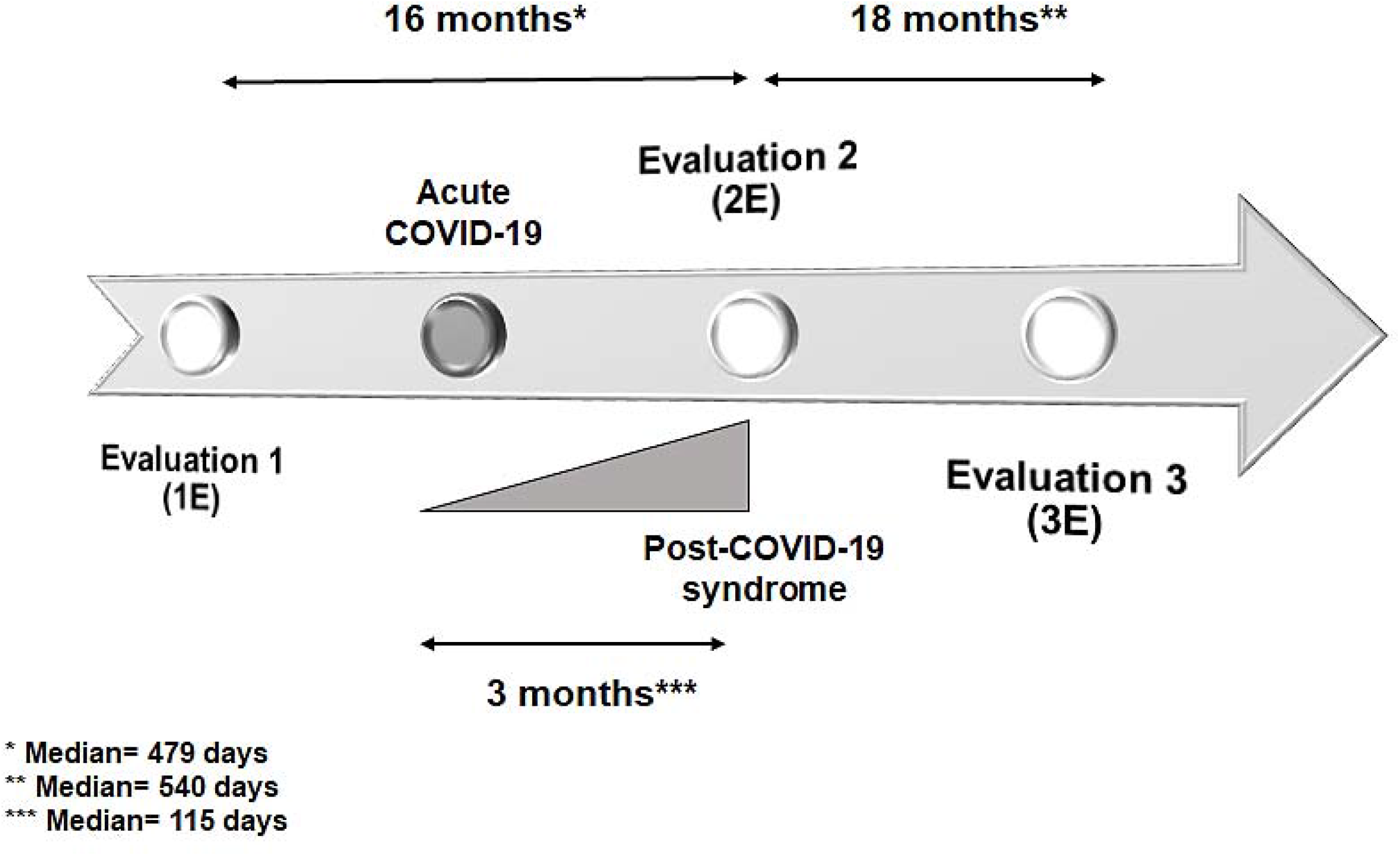
General scheme of the study

### Participants

The study was performed on the general population of a semi-urban area attended by a primary care center in northern Spain. Patients who suffered COVID-19 between April and September 2020 were selected. None of them had been vaccinated against SARS-CoV-2 at the time of inclusion, and all cases were managed exclusively in the primary care setting. SARS-CoV-2 infection was confirmed by a positive real-time reverse transcription-polymerase chain reaction (RT-PCR) test or by the presence of anti-SARS-CoV-2 IgG three months after acute COVID-19. A second inclusion criterion was a mild course of infection as defined by WHO [10] and characterized by fever, malaise, cough, upper respiratory symptoms and/or less common manifestations of COVID-19, in the absence of dyspnea. Previous diagnosis of T2DM and liver disease were considered exclusion criteria.

### Clinical variables and laboratory parameters

Epidemiological variables (sex, age, body mass index -BMI, measured in kg/m^2^-, smoking, Charlson comorbidity index -CCI-, comorbidities), clinical data in relation to the acute phase, the presence of a PCS, and laboratory tests, were analysed. Interviews were conducted by physicians from the research team, using a structured questionnaire [9].

The main study variable was the Triglyceride Glucose index (TyG index), a surrogate marker of IR, which presents a high correlation with the homeostatic model assessment of insulin resistance (HOMA-IR) [11]. The TyG index was evaluated as a quantitative variable with the formula *TyG index= log*_*n*_ *[plasma triglycerides (mg/dL) x basal blood glucose (mg/dL) / 2]*, and as a categorical variable, considering 2 cut-off points: 8.74 and 7.68, which were used to classify IR (≥8.74) and IS (<7.68), as commented in heading Operational Definitions. The median was used to distinguish between high (↑) and low (↓) values. TG/HDL index has also been analysed [12].

Albumin-to-Globulin ratio (AGR) and lactate dehydrogenase enzyme (LDH) have been evaluated as markers of inflammation. The AGR integrates nutritional and inflammatory status, and is defined by the albumin / (total protein - albumin) ratio. Widely used at present, it is considered a suitable biomarker of inflammation [13]. Inflammation can lead to a decrease in albumin and/or an increase in globulin (which includes interleukins, immunoglobulins and the complement system among others), and consequently to a lower AGR. In acute COVID-19, a decreased AGR has proven useful as an initial indicator of risk, as well as a prognostic factor for severity and mortality [14]. A cut-off point of 1.50 was selected, indicating inflammation below that figure [15].

LDH, the last enzyme of the glycolytic and lactate-generating pathway, is a clinically useful inflammatory marker. Elevated serum LDH levels have been associated with worse outcome in patients with various malignancies and in viral infections such as COVID-19 [16]. A serum LDH level of 206 U/L, the cut-off point for the third tertile at baseline, was used to determine the frequency of elevated serum LDH at 2E (LDH2) and at 3E (LDH3). Normality range was 120-246 U/L. Neutrophil-to-lymphocyte ratio (NLR) has also been evaluated.

### Operational definitions

Three grades of acute COVID episode intensity have been distinguished for illustrative purposes. Grade I corresponded to an episode with a number of symptoms below the median together with a duration -in days-also below the median; grade III was defined by figures of both variables above the median, and the remaining two situations were classified as grade II.

The diagnosis of PCS was established when the National Institute for Health and Care Excellence (NICE) criteria were met (signs and symptoms developing during or after an infection consistent with COVID-19, beyond 12 weeks of the acute episode, and not explained by an alternative diagnosis [17].

No single TyG index cut-off point has been defined to establish the diagnosis of IR and there is a wide variability across studies [18,19]. To that end, and in line with other authors [20], we selected the group with baseline plasma TG levels in the highest tertile and HDL in the lowest. The median TyG1 index was calculated in this group, and thus, when applied on TyG2 and TyG3, any value >8.74 was classified as IR. This figure matches with published studies, in which <8 is considered normal and 8.74 falls within the range of increased CV risk [21]. Similarly, we analysed the persons who presented a basal plasma TG level in the lowest tertile and HDL in the highest one, and the median of TyG was used to classify IS. Thus, TyG2 or TyG3 values <7.68 were considered IS.

The identification in the sample of basal CVD strongly associated with IR, such as obesity or hypertension, did not allow us to prove that COVID-19 generates new IR. To overcome this methodological drawback, a group of subjects without metabolic or CVD was identified and evaluated, and this procedure reasonably ruled out the bias of an adverse metabolic profile favoring IR. The following baseline criteria were used to constitute the subsample: BMI <25 kg/m^2^, absence of hypertension, dyslipidemia, chronic kidney disease or ischemic heart disease, AGR value >1.50 and serum LDH value below the median. T2DM were previously discarded in the current study, as commented.

Diagnoses of T2DM, dyslipidemia, hypertension and other baseline comorbidities were extracted from the clinical history and were based on clinical guidelines and international standards. Blood samples were obtained from an antecubital vein using the standard venipuncture procedure, in the morning and after a 12-hour fast.

Results were provided by our reference, tertiary-level hospital. Hematologic cell counts were analysed on a DXH900 (Beckman Coulter). Serum concentrations of glycemia, albumin and lipid profile were obtained by automated methods in an ADVIA 2400 Chemistry System autoanalyzer (Siemens, Germany). LDH was analysed by spectrophotometric assay on an Atellica CH analyzer (Siemens Healthcare Diagnostics Inc, Tarrytown, NY, USA).

### Statistical analysis

#### Missing data

An automatic multiple imputation process has been performed, a procedure that provides unbiased estimates, preserves sample size and allows statistical power to be maintained [22]. For this purpose, a previous missing values analysis was performed, in which all variables involved in the main comparisons were included, as well as others significantly correlated with the incomplete variables. Using the regression method, missing values were replaced by a random sample of plausible value imputations. Finally, the internal consistency of the procedure was checked with an analysis on original and imputed data.

#### Statistical methods

The Shapiro-Wilk test identified that quantitative variables deviated from the normal distribution and were expressed as median [interquartilic range (IQR)]. If a median was 0, the mean was used instead. Some quantitative variables have also been expressed as categorical variables using the median or tertiles, considering high (↑) or low (↓) levels above or below the median, respectively. Contrasts were performed using non-parametric tests, such as Spearman correlation. When subjects were compared with themselves at another point in time, repeated measures tests such as Friedman’s test or Wilcoxon signed-rank test were used.

Categorical variables were expressed as percentages, and statistical tests for contingency tables, such as Pearson’s chi-square or Fisher’s exact test, and McNemar’s test in the case of related samples, were used for comparisons.

Multiple linear regressions combined with stratified analysis were performed, and according to the objective of the study, addressing both sexes separately. In all cases, the dependent variable (DV) was the TyG3 index. Baseline variables that showed a significant association with TyG in the bivariate analysis were included in the model. Variables of interest for the research were also included, seeking a good adjustment without causing overadjustment. The models were validated by checking the requirement of normality of the standardized residuals and the Z-type curve approximation ∼ N(0,1).

Bonferroni correction for multiple comparisons was applied and a p-value of p<0.05 was considered significant in all calculations. The contrast between correlation coefficients was performed using a freeware from the web page https://www.psychometrica.de/korrelation.html, and the rest of the analyses were carried out with the IBM SPSS 25 statistical package (IBM Corp, Armonk, NY, USA).

#### Ethical considerations

The tenets of the Declaration of Helsinki in research on human subjects were followed. All patients who met the inclusion criteria were informed of the purpose of the study and were invited to participate. All of them expressed their verbal consent and there was no refusal to participate. The study was approved by the Clinical Research Ethics Committee of Cantabria (code 2021.102).

## RESULTS

### Baseline data

Initially, 131 patients with a confirmed COVID-19 were collected. Of these, 13 were excluded due to moderate or severe disease and 6 due to a previous diagnosis of T2DM. Thus, 112 subjects with a previous mild COVID-19 were finally included. In an initial phase, missing data analysis was performed followed by multiple data imputation by regression.

The sample was composed of young adults (69.8% were <53 years), with a similar frequency of both sexes, low comorbidity, and dyslipidemia and hypertension as the most frequent pathologies. A total of 14.3% of the participants was obese (**Table 1**). Median [IQR] of BG1 and TyG1 index were 87 [73-101] mg/dL and 8.25 [7.4-9.0], respectively (**Table 2**). TyG index scores ≥8.74 and <7.68 were considered IR and IS respectively, as commented.

**Table 1:** Clinical characteristics of the participants

|  | **TOTAL SAMPLE**<br>**(n=112)** |
| --- | --- |
| Females; *n*(%) | 65 (58) |
| Males; *n*(%) | 47 (42) |
| Age (years) | 44 [24-64] |
| Body mass index (kg/m<sup>2</sup>) | 24 [19-29] |
| Obesity; *n*(%) | 16 (14.3) |
| Tobacco; *n*(%) | 38 (33.9) |
| Hypertension; *n*(%) | 19 (17) |
| Dyslipidemia; *n*(%) | 36 (32.1) |
| Ischemic heart disease; *n*(%) | 4 (3.6) |
| Cerebrovascular disease; *n*(%) | 1 (0.9) |
| Immunosuppression; *n*(%) | 3 (2.7) |
| Asthma; *n*(%) | 11 (9.8) |

*Quantitative variables, expressed as median [interquartile range]*

**Table 2:** Glycemic parameters and inflammatory markers at data points, in different strata

|  | TOTAL SAMPLE |  |  |  | FEMALES |  |  |  | MALES |  |  |  |
| --- | --- | --- | --- | --- | --- | --- | --- | --- | --- | --- | --- | --- |
|  | 1E | 2E | 3E | p* | 1E | 2E | 3E | p* | 1E | 2E | 3E | p* |
| Basal glycemia (mg/dL) | 87 [14] | 90 [11] | 89 [14] | **0.014** | 86 [13] | 88 [10] | 88 [15] | **0.019** | 86 [12] | 89 [11] | 90 [15] | 0.15 |
| TyG index | 8.25 [0.8] | 8.31 [0.7] | 8.32 [0.7] | **0.002** | 8.10 [0.9] | 8.24 [0.8] | 8.39 [0.6] | **0.004** | 8.26 [0.7] | 8.29 [0.7] | 8.26 [0.7] | 0.20 |
| TyG index >8.74; n(%) | 23 (20.5) | 29 (25.9) | 31 (27.7) | 0.23 | 15 (23.1) | 17 (26.2) | 19 (29.2) | 0.48 | 8 (17) | 12 (25.5) | 12 (25.5) | 0.45 |
| TG/HDL | 1.47 [1] | 1.76 [2] | 1.68 [1] | **0.049** | 1.39 [1] | 1.42 [1] | 1.67 [1] | 0.06 | 1.73 [2] | 1.70 [2] | 1.75 [1] | 0.43 |
| Albumin-to-Globulin ratio | 1.82 [0.5] | 1.55 [0.3] | 1.77 [0.3] | 0.66 | 1.75 [0.5] | 1.53 [0.2] | 1.79 [0.3] | 0.23 | 1.85 [0.4] | 1.61 [0.3] | 1.72 [0.5] | 0.51 |
| AGR <1.50; n(%) | 25 (22.3) | 40 (35.7) | 10 (8.9) | **0.012** | 16 (24.6) | 25 (38.5) | 4 (6.2) | **0.012** | 9 (19.1) | 15 (31.9) | 6 (12.5) | 0.54 |
| Lactate dehydrogenase (U/L) | 176 [44] | 183 [39] | 185 [54] | 0.35 | 177 [43] | 177 [42] | 185 [53] | 0.72 | 177 [48] | 181 [50] | 186 [74] | 0.28 |
| LDH >206 U/L; n(%) | 18 (16.1) | 24 (21.4) | 36 (32.1) | **0.007** | 10 (15.4) | 13 (20) | 19 (29.2) | 0.06 | 8 (17) | 11 (23.4) | 17 (36.2) | 0.07 |
|  | BMI > 25 Kg/m<sup>2</sup> |  |  |  | POST-COVID-19 SYNDROME |  |  |  | GRADES II-III ACUTE COVID-19 |  |  |  |
|  | 1E | 2E | 3E | p* | 1E | 2E | 3E | p* | 1E | 2E | 3E | p* |
| Basal glycemia (mg/dL) | 91 [15] | 91 [11] | 92 [13] | 0.07 | 85 [13] | 90 [9] | 85 [14] | 0.92 | 87 [11] | 90 [11] | 87 [11] | **0.016** |
| TyG index | 8.48 [0.5] | 8.64 [0.6] | 8.58 [0.5] | 0.08 | 8.27 [0.9] | 8.31 [0.8] | 8.33 [0.8] | 0.88 | 8.12 [0.8] | 8.30 [0.8] | 8.33 [0.7] | **0.007** |
| TyG index >8.74; n(%) | 16 (29.1) | 24 (43.6) | 19 (34.5) | 0.35 | 11 (30.6) | 10 (27.8) | 12 (33.3) | **0.011** | 19 (22.6) | 25 (29.8) | 26 (31) | 0.07 |
| TG/HDL | 1.47 [1] | 1.76 [2] | 1.68 [1] | 0.42 | 1.64 [1] | 1.67 [2] | 1.60 [1] | 0.69 | 1.44 (1) | 1.96 (2) | 1.78 (1) | **0.014** |
| Albumin-to-Globulin ratio | 1.82 [0.5] | 1.55 [0.3] | 1.77 [0.3] | 0.28 | 1.75 [0.5] | 1.49 [0.4] | 1.76 [0.3] | 0.70 | 1.83 [05] | 1.61 [0.3] | 1.79 [0.3] | 0.96 |
| AGR <1.50; n(%) | 12 (21.8) | 29 (52.7) | 5 (9.1) | 0.30 | 9 (25) | 17 (47.2) | 6 (16.8) | 0.60 | 19 (22.6) | 21 (32.1) | 8 (9.3) | 0.86 |
| Lactate dehydrogenase (U/L) | 176 [44] | 183 [39] | 185 [54] | 0.09 | 184 [48] | 183 [49] | 185 [70] | 0.76 | 178 [45] | 180 [37] | 180 [52] | 0.69 |
| LDH >206 U/L; n(%) | 11 (20) | 16 (29.1) | 20 (36.4) | 0.98 | 5 (13.9) | 7 (19.4) | 12 (33.3) | 0.17 | 15 (17.8) | 18 (21.4) | 27 (32.1) | 0.47 |

Quantitative variables, expressed as median [interquartile range]
p\*: Contrast between 1E and 3E; TyG index: Triglyceride Glucose index
1E: Baseline assessment; 2E: Assessment 3 months after acute COVID-19; 3E: Assessment 20 months after acute COVID-19.

LDH1 was found to be positively associated with number of symptoms of acute COVID-19 (0.83 ± 0.1 vs 0.43 ± 0.1; p=0.049) and with TyG1 index (Serum LDH >206 U/L in 36.8% of subjects with ↑TyG1 index vs 14.1% of those with ↓TyG index; p=0.022). In addition, LDH1 and TyG3 index were significantly correlated in both sexes (**Table 3**), and this relation was influenced by PCS (**Figure 2**).

**Table 3:** Significant correlations between TyG index and covariates of interest, stratifying by sex

|  |  | FEMALES (n=65) |  |  |  |  |  |  |  |  |  |  |
| --- | --- | --- | --- | --- | --- | --- | --- | --- | --- | --- | --- | --- |
|  |  | TyG3 index | Age (years) | BMI | CCI | AGR1 | LDH1 | Acute COVID: symptoms | Acute COVID: days | PCS: symptoms | AGR2 | LDH2 |
| MALES (n=47) | TyG3 index |  |  | 0.359** |  |  | 0.214* |  |  |  |  |  |
|  | Age (years) |  |  | 0.333** |  | 0.311* |  |  |  |  |  | 0.294* |
|  | BMI |  |  |  |  |  |  |  |  |  | - 0.468** |  |
|  | CCI |  | 0.382** |  |  |  |  |  |  |  |  |  |
|  | AGR1 |  |  |  |  |  | 0.246* |  |  |  | 0.257* | 0.250* |
|  | LDH1 | 0.298* | 0.319* | 0.311* |  |  |  |  |  |  |  | 0.491* |
|  | Acute COVID: symptoms |  |  |  |  |  |  |  | 0.539** | 0.500** |  |  |
|  | Acute COVID: days |  | 0.421** |  |  |  |  | 0.404** |  | 0.321* |  |  |
|  | PCS: symptoms |  |  |  |  | - 0.362* |  |  |  |  |  |  |
|  | AGR2 |  |  | - 0.292* | - 0.403** | 0.292* |  |  |  |  |  |  |
|  | LDH2 |  |  | 0.335* |  |  |  |  |  |  |  |  |

*TyG index: Triglyceride glucose index; BMI: Body mass index; CCI: Charlson comorbidity index; AGR: Albumin-to-globulin ratio; LDH: Lactate dehydrogenase;*
*PCS: Post-COVID-19 syndrome.*
*\*p <0.05; \*\*p <0.01*

**Figure 2:**
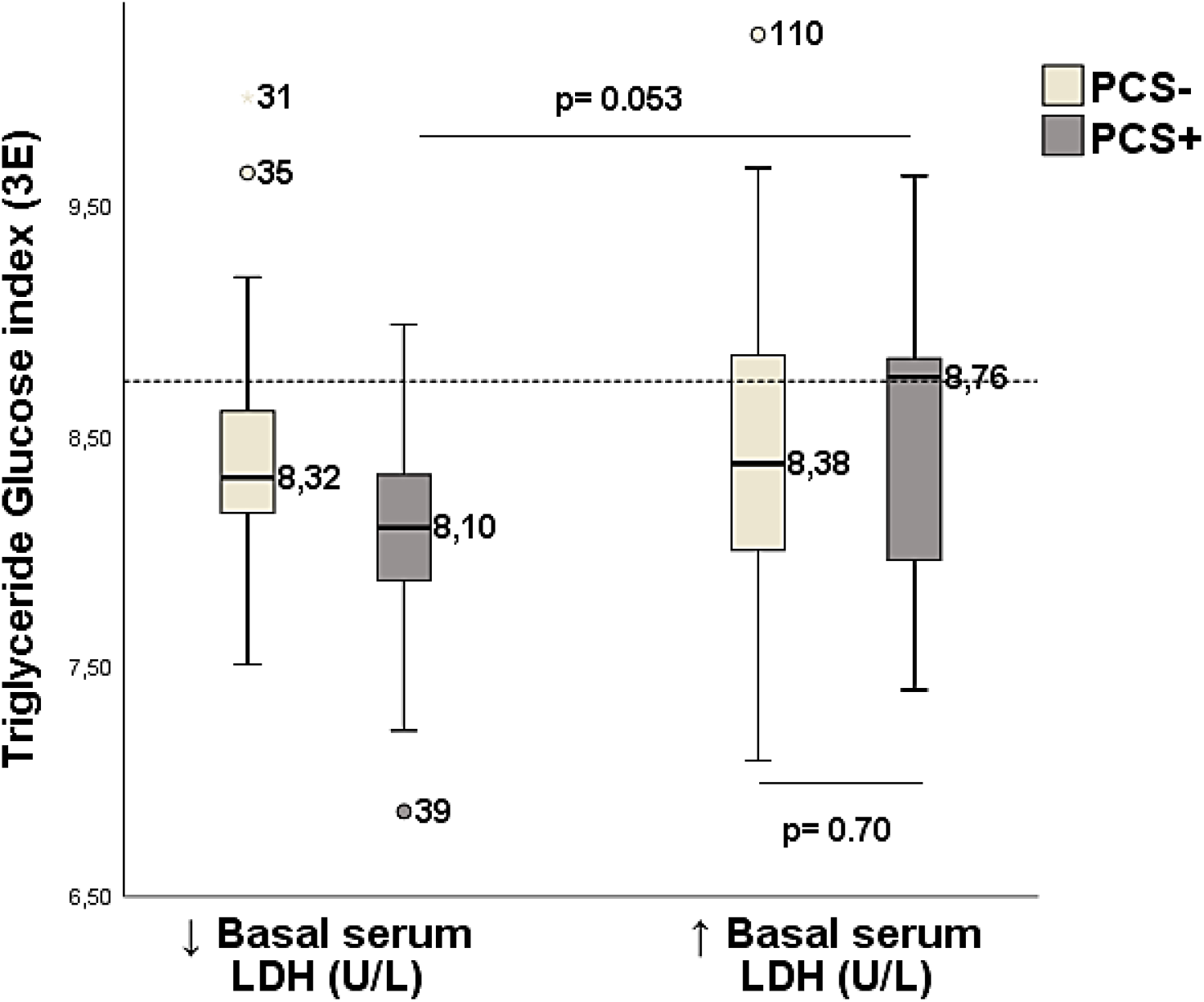
Triglyceride Glucose index, in relation to basal serum LDH level and post-COVID-19 syndrome *PCS: Post-COVID-19 syndrome. LDH: lactate dehydrogenase* *Dashed line at 8.74 represents the TyG index score for classifying insulin resistance*

In acute COVID-19, the most frequent symptoms were fever (60.3%), anosmia/ageusia (52.9%), cough (46.3%), fatigue (43.8%), myalgia (46.3%), headache (42.1%), odynophagia (32.2%), rhinitis (21.5%) and dyspnea (17.4%), with medians of 5 [1-9] symptoms and 10 [1-19] days. Grade I symptoms occurred in 21.5%, grade II in 33.6%, and grade III in 44.8% of patients.

Thirty-six subjects, 31.3% of the sample, had PCS. Of these, 26 were women and 10 were men (prevalences of 40% and 21.3%, respectively; p=0.036). The most frequent symptoms of PCS were fatigue (42.8%), anosmia (40%), ageusia (22.8%), dyspnea (17.1%), myalgia (11.4%) and palpitations (11.4%).

### The three data points

As shown in **Table 2**, significant differences were recorded between 1E and 3E. While the prevalence of IR was 20.5%, 25.9% and 27.7% at 1E, 2E and 3E, respectively, prevalence of IS was 20.5% (1E), 9.8% (2E) and 7.1% (3E).

A total of 8 previously normoglycemic subjects, had BG2 (7 of them) or BG3 (1 participant) >126 mg/dL. Considering the total sample, BG, TyG index and TG/HDL raised after acute COVID-19, and remained elevated at 3E, with significant differences with respect to 1E. In parallel, the percentage of subjects with high LDH levels increased at 2E and 3E. AGR presented at 2E the lowest value and the highest frequency of values <1.50, compared to 1E and 3E.

These patterns (increased BG, IR and LDH levels with respect to baseline values, and decreased AGR figures at 2E) were also observed in both sexes, although the differences only reached significance in women. TyG index value of 8.74 has identified the groups with IR highest prevalence at 3E: subjects with BMI ≥25 kg/m^2^ and PCS+ individuals (**Figure 3**). Likewise, both groups also showed the highest frequency of AGR <1.50 (52.7% and 47.2%, respectively), while subjects belonging to the groups BMI <25 kg/m^2^, PCS-, and males, displayed a prevalence of AGR <1.50 ranging between 19.3% and 31.9%.

**Figure 3:**
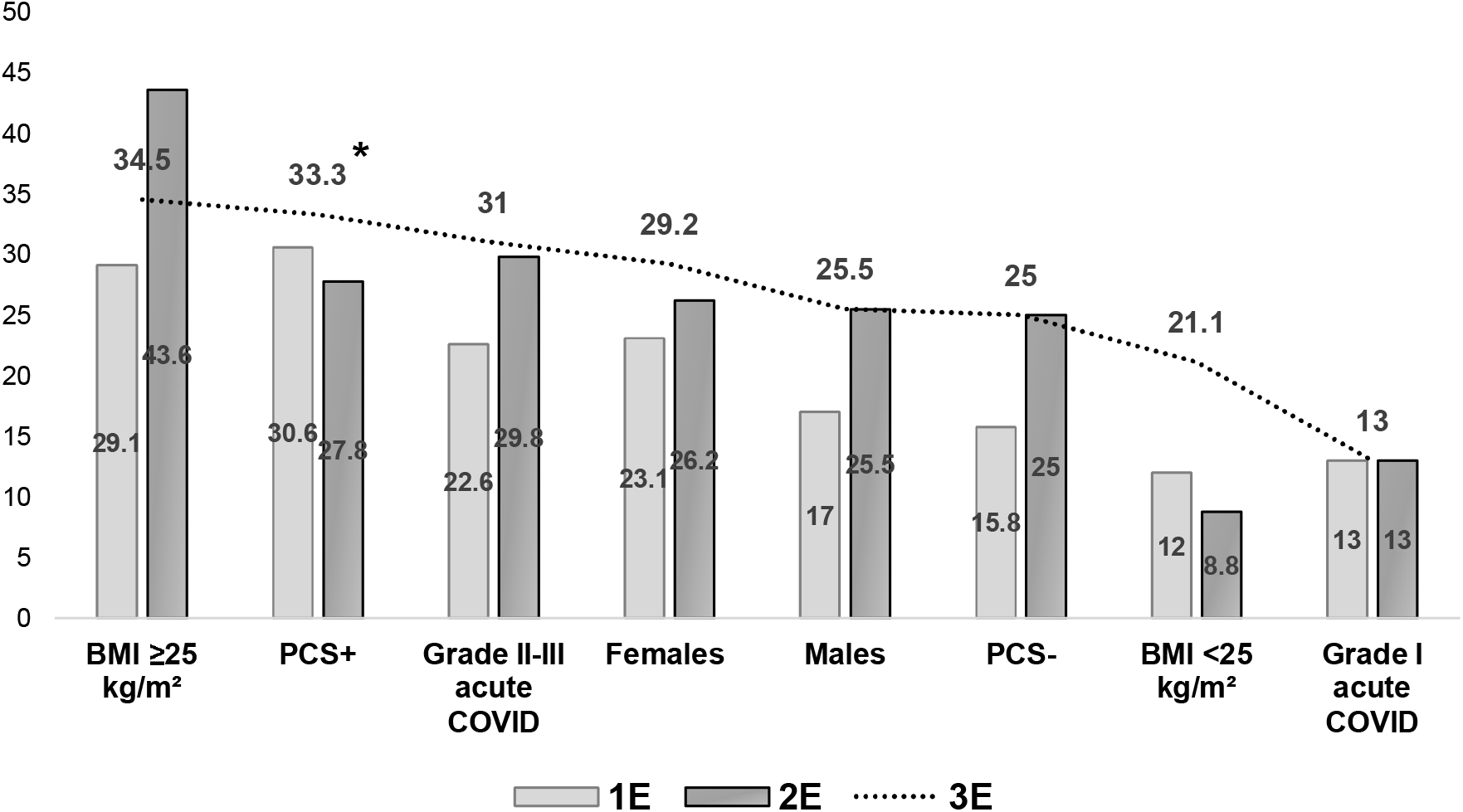
Frequency of insulin resistance at data points *BMI: Body mass index; PCS: Post-COVID-19 syndrome* *\* p<0.05 (contrast between 3E and 1E)*

### The group without metabolic or CV disease

As mentioned, this group was composed of 15 subjects of both sexes, without metabolic disorder or CV disease. Subjects in this group were younger, with normal weight and had lower baseline values of serum LDH and TyG index (**Table 4**). They also had lower plasma TG levels (68 [48-88] vs 76 [61-91]; p=0.039). The intensity of acute COVID-19 and the prevalence of PCS did not differ significantly from the rest of the sample. One of them, a previously normoglycemic woman with BMI <25 kg/m^2^, presented with BG2=147 mg/dL.

**Table 4:**
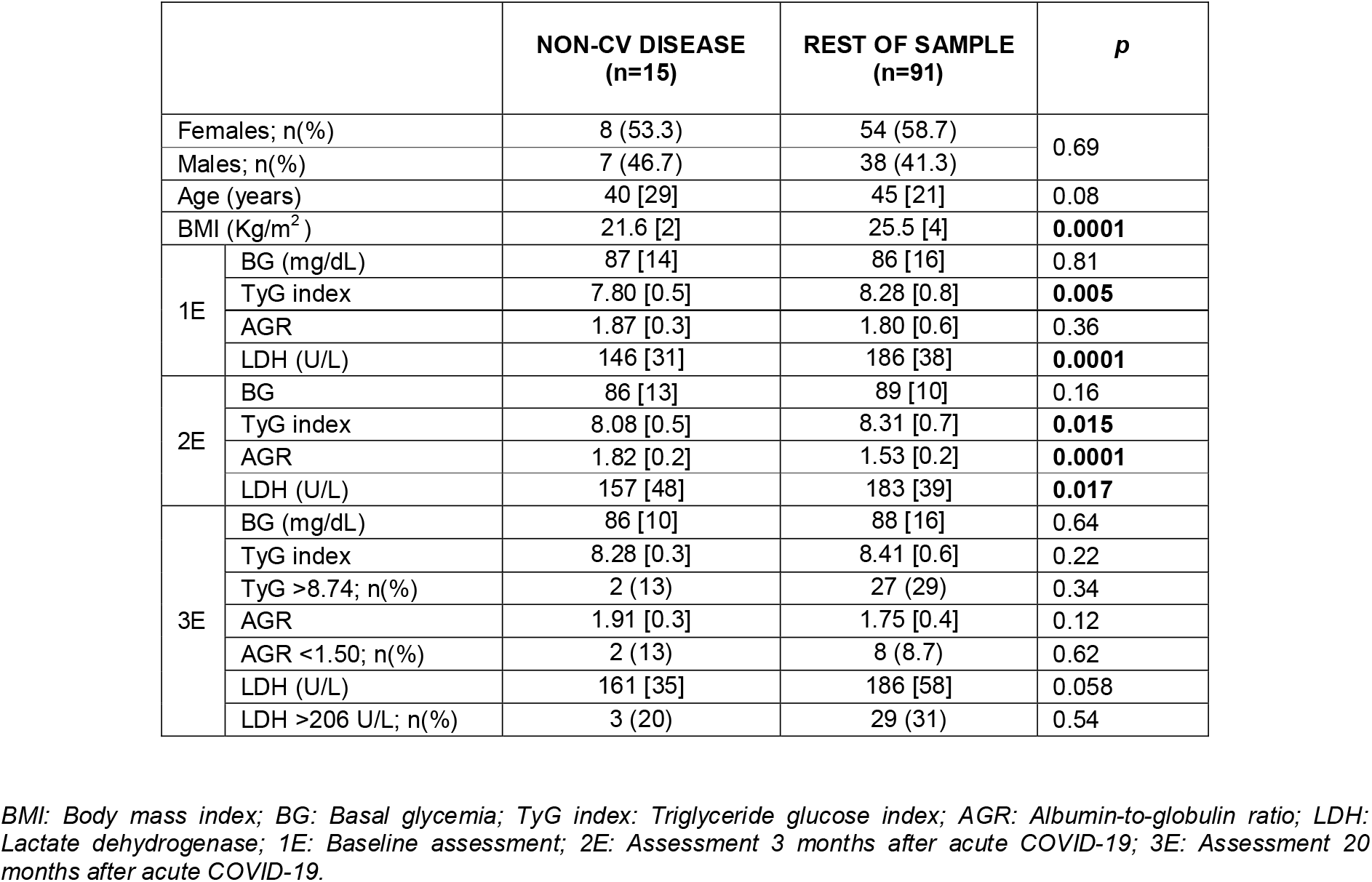
Clinical characteristics of the group without metabolic or cardiovascular disease

This group showed a significant increase in the TyG index (TyG1 index= 7.80 [7.7-7.8], and TyG3 index= 8.28 [8.2-8.3]; p=0.017), and in the percentage of IR subjects (from 6.7% to 13.3%; p=0.11). Frequencies of IS individuals were 33.3% (1E), 20% (2E) and 6.7% (3E)

As a consequence, the significant differences compared to the rest of the sample observed at 1E and 2E, disappeared at 3E. Further analysis in this group revealed that the persons who had higher TyG3 index presented with higher NLR2 (2.40 [1.5-3.3] vs 1.52 [0.8-2.2]; p= 0.036). In this group, LDH1 and TyG3 were not correlated. In contrast, both variables were positively correlated in the rest of the sample (rho=0.211; p=0.043).

A total of 13 subjects (86.7%) belonging to this group had a grade II-III acute COVID-19, and they presented significantly higher serum LDH2 levels (161 [138-184] U/L) and TyG2 index values (8.29 [7.9-8.3]).

### Acute COVID-19, post-COVID-19 syndrome, and glycemic parameters

Together with those with BMI ≥25 Kg/m^2^, PCS+ subjects presented an AGR2 value <1.50 (**Table 2**) in the total sample and in both sexes (female PCS+, AGR=1.49 [0.7-2.1]; male SPC+, AGR=1.49 [0.8-2.2]). In addition, the individuals with the two characteristics, baseline BMI ≥25 kg/m^2^ and affected by PCS, displayed an AGR2 value of 1.44 [1.2-1.6].

In females, PCS was associated with intensity of acute COVID-19, while in men, PCS was associated with basal inflammation: Up to 80% of PCS+ women had an acute episode classified as grade II-III, versus 37.8% of PCS-women (p=0.005). In contrast, PCS affecting males was associated with AGR1 and TyG1 index: PCS+ males showed lower AGR1 (62.5% with AGR1 in the lower tertile vs. 16.1% in PCS-males; p=0.012) and a high prevalence of IR (40% with TyG1 index >8.74 vs. 10.8% in PCS-males; p=0.029).

PCS has shown influence on the variations of current IR, enhancing the associations between BMI and TyG3, and between LDH1 and TyG3. Thus, the correlation between BMI and TyG3 index was increased by the presence of PCS, from r= 0.158 (p=0.13) in PCS-to r= 0.497 (p=0.002) in PCS+. Also, by a grade II-III acute COVID-19, from r=0.058 (p=0.79) in grade I to r= 0.383 (p=0.0001) in grade II-III. On the other hand, a subject with LDH1 >178 U/L and PCS, compared to those with low LDH1 and PCS, showed a higher TyG3 index value (p=0.053), in range of IR (**Figure 2**)

### Spearman correlation

As shown in **Table 3**, two baseline covariates, BMI and LDH1, were significantly correlated with TyG3 index. Likewise, these factors gathered most of the significant correlations. There were relevant differences by sex in the intensity of the different correlations involving PCS: In women, the number of PCS symptoms strongly correlated with the number of symptoms in acute COVID-19, with rho=0.500 (p=0.0001); in contrast, in men, rho=0.059 (p=0.67). The difference between both coefficients was significant (p=0.001). In males, AGR1 was inversely correlated with the number of PCS symptoms (rho= -0.362; p=0.012), while in females there was no correlation, with rho=0.071; p=0.57 (p=0.011 for the difference between both coefficients).

In women, plasma fibrinogen level at 2E (FBG2) correlated significantly with TyG2 (rho=0.330; p=0.009) and TyG3 (rho=0.271; p=0.033). In contrast, both correlations were weaker and non-significant in men.

Spearman correlation between TyG3 and LDH1, LDH2 and LDH3, showed rho values of 0.232 (p=0.014), 0.162 (p=0.087) and -0.059 (p=0.53), respectively.

### Multivariable analysis

Considering the aim of the study and the results of bivariable analysis, we have built several multiple linear regressions addressing the different combinations of BMI and both sexes. A total of 8 models, all of them with TyG3 as the DV, and the same adjustment variables (**Table 5**).

**Table 5:**
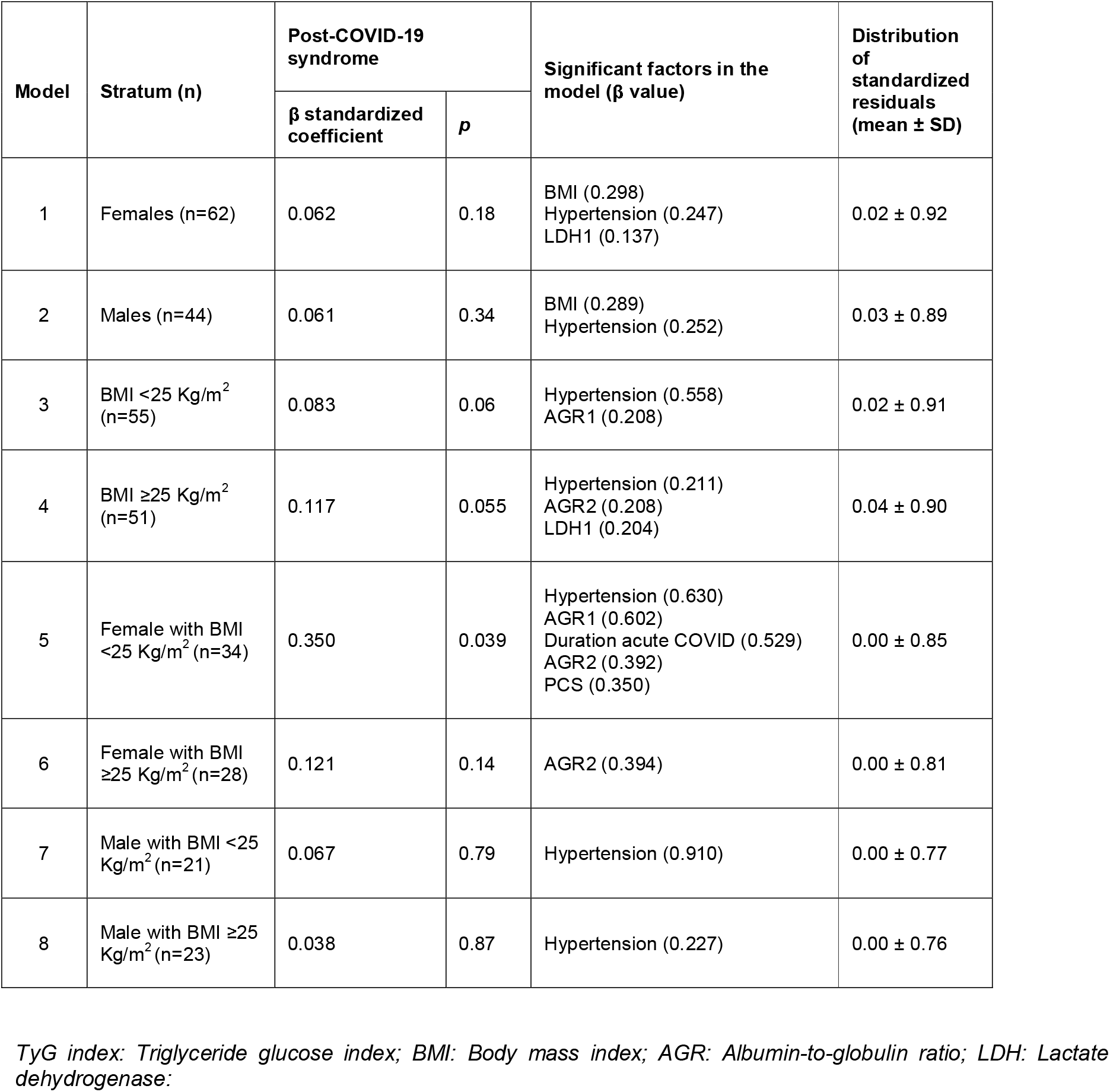
Multivariable models including post-COVID-19 syndrome as independent variable, with current TyG index as output variable

| Model | Stratum (n) | Post-COVID-19 syndrome | | Significant factors in the model ($\beta$ value) | Distribution of standardized residuals (mean ± SD) |
| --- | --- | --- | --- | --- | --- |
| | | $\beta$ standardized coefficient | $p$ | | |
| 1 | Females (n=62) | 0.062 | 0.18 | BMI (0.298)<br>Hypertension (0.247)<br>LDH1 (0.137) | 0.02 ± 0.92 |
| 2 | Males (n=44) | 0.061 | 0.34 | BMI (0.289)<br>Hypertension (0.252) | 0.03 ± 0.89 |
| 3 | BMI <25 Kg/m<sup>2</sup> (n=55) | 0.083 | 0.06 | Hypertension (0.558)<br>AGR1 (0.208) | 0.02 ± 0.91 |
| 4 | BMI ≥25 Kg/m<sup>2</sup> (n=51) | 0.117 | 0.055 | Hypertension (0.211)<br>AGR2 (0.208)<br>LDH1 (0.204) | 0.04 ± 0.90 |
| 5 | Female with BMI <25 Kg/m<sup>2</sup> (n=34) | 0.350 | 0.039 | Hypertension (0.630)<br>AGR1 (0.602)<br>Duration acute COVID (0.529)<br>AGR2 (0.392)<br>PCS (0.350) | 0.00 ± 0.85 |
| 6 | Female with BMI ≥25 Kg/m<sup>2</sup> (n=28) | 0.121 | 0.14 | AGR2 (0.394) | 0.00 ± 0.81 |
| 7 | Male with BMI <25 Kg/m<sup>2</sup> (n=21) | 0.067 | 0.79 | Hypertension (0.910) | 0.00 ± 0.77 |
| 8 | Male with BMI ≥25 Kg/m<sup>2</sup> (n=23) | 0.038 | 0.87 | Hypertension (0.227) | 0.00 ± 0.76 |

*TyG index: Triglyceride glucose index; BMI: Body mass index; AGR: Albumin-to-globulin ratio; LDH: Lactate dehydrogenase.*

PCS has shown to be an independent and predictive variable of TyG in women with BMI <25 kg/m^2^, after controlling for age, hypertension, BMI, Charlson comorbidity index, AGR1, AGR2, LDH1, number of symptoms of acute COVID-19, and number of days of the acute episode. In this model, hypertension (β=0.630), AGR1 (β=0.602), duration of the acute COVID-19 (β=0.529), AGR2 (β=0.392) and PCS (β=0.350) maintained statistical significance after adjustment.

## DISCUSSION

The present study has demonstrated progressive increases of BG, serum LDH and TyG index along the data points. Likewise, it has also shown remarkable sex differences and findings of interest in three lines: (i) grade II-III acute COVID-19 and PCS have acted similarly with respect to IR, (ii) data supporting the theory of COVID-induced new IR, and (iii) a close relationship observed between LDH1 and TyG3. These findings are discussed below.

### Grade II-III acute COVID-19 and post-COVID-19 syndrome

It is pertinent to note the close relations observed between PCS and grade II-III acute COVID-19: Both groups, along with those with BMI >25 Kg/m^2^, starting from high baseline levels, reached the highest TyG figures. In women, the number of PCS symptoms was correlated with duration and number of symptoms of acute COVID-19. Moreover, PCS and acute COVID-19 reinforced the relationship between BMI and TyG index in a similar way. Regarding the models analysed, both have been shown to be consistent predictors of the TyG3 index in the same model, the one aimed at women with BMI <25 kg/m^2^.

PCS and acute grade II-III COVID-19 have acted analogously, and we conjecture that they are two facets of the same disorder: a persistent low-grade inflammatory state with incremental effect on IR.

An intense acute COVID-19 seems to act as a trigger regarding PCS [9] and the glucose metabolism impairment [6].

Viral infections activate the inflammasome and stimulate production of cytokines/chemokines. Of them, IFNγ, IL-1β, IL-6 and TNFα, whose levels have been found to be persistently elevated in the PCS, have a deleterious effect of increased IR [23,24]. Thus, IFNγ production by NK cells in skeletal muscle is stimulated by viral infection and induces transcriptional down-regulation of the insulin receptor in myocytes, resulting in systemic IR [25]. Pancreatic β-cells express the receptor for IL-1β. Infections elevate circulating IL-1β, and chronically elevated levels can cause β-cell dysfunction [26]. Similarly, persistently elevated levels of IL-6 induce apoptosis in pancreatic islets along with other inflammatory cytokines and contribute to IR [27]. In turn, TNFα stimulates the cells to produce more lactate [28] and overproduction of TNFα in adipose tissue appears to feed inflammation, β-cell death and produces additional IR in peripheral tissues [29].

Multivariate analysis has pointed to PCS as a significant predictor of IR in normal weight women. Interestingly, the association was nullified when plasma FBG2 was added in the model. This finding agrees with the close relationship previously observed in women between FBG2 and PCS [9], and in the current study, also in women, between FBG2 and TyG2/TyG3.

Nevertheless, in terms of predicting IR, PCS has played a secondary role compared to BMI or hypertension, which have been shown to be much relevant predictors in both sexes. It is known that hypertension and obesity present strong links with IR, with chronic low-grade inflammation and oxidative stress as underlying mechanisms, and a substantial increase in the risk of developing T2DM and CVD [30,31].

### The subgroup without CVD and IR *de novo*

SARS-CoV-2 infection has been said to push patients with prediabetes toward T2DM, and patients with undiagnosed T2DM toward severe complications [32]. In the study, 14.3% was obese and 17% was classified as hypertensive, accounting for a certain degree of baseline IR. In this context, the group without metabolic disturbances -composed of younger individuals, with BMI <25 kg/m^2^, TyG1 index <8 and without markers suggestive of inflammation-ruled out prior IR and acted as a control group. Therefore, the result of a significant increase in IR can be considered a new IR.

As commented, cohort studies have reported a significant increase in the incidence of T2DM according to the severity of the acute phase [6,33]. Up to 86% of persons in this group had a grade II-III acute COVID-19, a feature associated in the study with significant elevations of plasma BG and TyG index. In addition, subjects with a higher TyG3 index showed a high NLR2. These findings could corroborate that an intense acute COVID-19 promotes an inflammatory milieu and metabolic alterations leading to new IR in patients with no previous apparent IR.

### The relationship between baseline LDH and current TyG

LDH is an enzyme present in almost every cell of the body and converts pyruvate to lactate in the glycolytic pathway under conditions of oxygen insufficiency. Elevation of LDH is a sensitive indicator of increased cell membrane permeability and cell injury as well. LDH is a metabolic marker of physiological distress, often used to diagnose myocardial infarction, vessel damage, tissue injury, and certain types of malignant tumors [34].

We found increasing levels of serum LDH throughout the 3 assessments, and a consistent association between LDH1 and TyG3 index. In addition to being significantly correlated in both sexes, LDH1 was a consistent predictor of TyG3 index in two multivariate models, females and overweight/obese individuals. Unexpectedly, serum LDH2 and LDH3 showed no such association.

Several mechanisms accounting for the relationship between LDH1 and TyG3 index could be speculated, in particular specific associations of both parameters with MetS. MetS is a condition of chronic low-grade inflammation as a consequence of complex interplay between genetic and environmental factors, in which IR seems to exert a key role [35]. Studies have reported high LDH levels in association with MetS [36,37] or with its components, such as hypertension, where high LDH levels have been related to albuminuria and glomerular endothelial damage [38] or T2DM, with a direct relation between LDH levels and BG, and increased LDH in association with short-term glycemic variability [34]. Certain hormones and growth factors, such as insulin and insulin-like growth factor-1 (IGF-1), stimulate the tissues to produce more lactate [28]. In this line, lactate is elevated in hyperinsulinemic subjects with normal fasting glucose, a situation frequently observed at the onset of IR [39,40]. It has been said that lactate is not only increased in the early stages of T2DM, but has also been shown to be predictive of its onset in the future [39].

In the same line, TyG index is also associated with MetS-related conditions including obesity, diabetes and hypertension [41,42] and predicts cardio-cerebrovascular disease [42,43]. In addition, both LDH as well as TyG, have been shown to be correlated with high-sensitive CRP [44,45] which is in turn closely related to CVD [46]. These associations involving LDH and TyG are likely mediated by miscellaneous connections between endothelial dysfunction, oxidative stress, IR and inflammation [36,42]. In the study, baseline clinical characteristics of some participants (14% obese, 17% hypertensive, 32% DLP, with TyG1 between 8.44 and 8.60 in these groups) carried a burden of IR that could explain the close interconnection between LDH1 and TyG3. It is worth mentioning that in the group without CV or metabolic disease, LDH1 did not show such an association, which could indirectly confirm our conjecture.

At this point, it is challenging to explain why only LDH1 showed a significant relationship with TyG3. Since LDH2 and LDH3 were generated after SARS-CoV-2 infection, one could speculate that they may present some differences respect to basal LDH. In this rationale, the difference could be driven by the isoenzyme profile.

LDH isoenzyme 1 (Iso1) and iso2 predominate in cardiac muscle, erythrocytes and kidneys. In contrast, Iso4 and Iso5 are dominant in liver and skeletal muscle. Iso3 is predominantly expressed in lymphoid tissue, brain, platelets and many malignant tissues. Its expression is also observed during lung infections. Although individual LDH isoenzymes tend to predominate in certain tissues, most tissues produce all LDH isoenzymes. Therefore, even in tissue-specific lesions, all isoenzyme levels increase with the expected predominant isoenzyme [28]. In acute SARS-CoV-2 infection, increased total serum LDH and iso3 levels, both associated with worse outcomes, have been described [16,47]. It has also been stated that COVID-19 patients with elevated LDH levels tended to experience PCS [24]. Accordingly, in the study, we observed a significant association between LDH1 >246 U/L and PCS, but not between LDH2 or LDH3 >246 U/L, and PCS.

Along with PCS, other differences that would support the hypothesis of different activated isoenzymes were observed. Although not shown, we have noted that (i) subjects with ↑LDH1 showed significantly higher basal TyG and TG/HDL indices compared with ↓LDH1 subjects, (ii) ↑LDH2 subjects showed significantly lower AGR1, and (iii) ↑LDH3 individuals had significantly higher NLR3 and lower AGR3. In summary, before infection, elevated LDH1 was associated with IR parameters, and after infection, elevated LDH2 and LDH3 were associated with parameters with a more inflammatory profile, such as AGR and NLR, possibly with the involvement of different isoenzymes respect to basal serum LDH.

Some caveats are necessary when interpreting the results. The study was performed in patients from a Caucasian and semi-urban population in northern Spain, so the results cannot be extrapolated to other populations or geographic areas. Another limitation has been the unavailability of LDH isoforms, which would have allowed us to gain knowledge on the profile associated with insulin resistance in our patients. Finally, 3E was performed with 87% of the sample being vaccinated against SARS-CoV-2, and possible adverse reactions affecting glucose metabolism could potentially bias the results. A search on the WHO Collaborating Centre for International Drug Monitoring (http://www.vigiaccess.org/) for the number of reported COVID-19 vaccine-related adverse reactions pertaining to terms related to glucose metabolism or pancreatitis (accessed April 9, 2023), yielded the following results: “hyperglycemia”, 1943 notifications; “DM”, 1848; “T2DM”, 926; “diabetic ketoacidosis”, 616; “glucose intolerance”, 146; “pancreatitis”, 1144. Although an alteration of glucose metabolism is potentially possible, this bias seems rather unlikely.

A strong point of our study is that it provides details on the relationship between PCS and IR after mild COVID-19, to our knowledge little explored. The sample without CVD is also noteworthy, since it allowed us to learn that COVID-19 can induce IR de novo, in patients without apparent IR.

## CONCLUSIONS

PCS has been shown to influence variations in IR, reinforcing the associations between BMI and basal LDH, with the current TyG index. In terms of predicting IR, PCS has played a secondary role, presenting a modest effect compared to BMI or prior hypertension.

Twenty months after mild COVID-19, patients have shown a significant increase in IR, both in cases of previous baseline IR and in those without previous IR. Basal serum LDH has shown to be predictive of current TyG, regardless of elevated LDH after SARS-CoV-2 infection. There were profound differences between women and men, confirming the need for a sex-stratified analysis when addressing the relation between PCS and glycemic alterations.

Further studies are warranted to confirm the results.

## Data Availability

All data produced in the present study are available upon reasonable request to the authors

## ABBREVIATIONS

ACE2: Angiotensin converting enzyme-2
AGR: Albumin-to-globulin ratio
BG: Basal glycemia
BMI: Body mass index
CCI: Charlson comorbidity index
CRP: C-reactive protein
CVD: Cardiovascular disease
IF_L_: Interferon gamma
IGF-1: Insulin growth factor-1
IL-1β: Interleukin-1β
IL-6: Interleukin-6
IR: Insulin resistance
IS: Insulin sensitivity
LDH: Lactate dehydrogenase
MetS: Metabolic syndrome
NLR: Neutrofil-to-lymphocyte ratio
PCS: Post-COVID-19 syndrome
TNFα: Tumor necrosis factor Alpha
TyG index: Triglyceride glucose index
T2DM: Type 2-diabetes mellitus

---

STROBE Statement—Checklist of items that should be included in reports of ***cross-sectional studies***

|  | Item No | Recommendation | Page |
| --- | --- | --- | --- |
| Title and abstract | 1 | (a) Indicate the study's design with a commonly used term in the title or the abstract | 3 |
|  |  | (b) Provide in the abstract an informative and balanced summary of what was done and what was found | 3 |
| **Introduction** |  |  |  |
| Background/rationale | 2 | Explain the scientific background and rationale for the investigation being reported | 4 |
| Objectives | 3 | State specific objectives, including any prespecified hypotheses | 5 |
| **Methods** |  |  |  |
| Study design | 4 | Present key elements of study design early in the paper | 5 |
| Setting | 5 | Describe the setting, locations, and relevant dates, including periods of recruitment, exposure, follow-up, and data collection | 5-6 |
| Participants | 6 | (a) Give the eligibility criteria, and the sources and methods of selection of participants | 5 |
| Variables | 7 | Clearly define all outcomes, exposures, predictors, potential confounders, and effect modifiers. Give diagnostic criteria, if applicable | 6-8 |
| Data sources/measurement | 8* | For each variable of interest, give sources of data and details of methods of assessment (measurement). Describe comparability of assessment methods if there is more than one group | 6 |
| Bias | 9 | Describe any efforts to address potential sources of bias | 7 (Subsample without CVD)<br>8 (Analysis post-imputations, missing data)<br>9 (Multivariable analysis: adjustment for confounders) (Bonferroni correction) |
| Study size | 10 | Explain how the study size was arrived at | 5-6 |
| Quantitative variables | 11 | Explain how quantitative variables were handled in the analyses. If applicable, describe which groupings were chosen and why | 7-9 |
| Statistical methods | 12 | (a) Describe all statistical methods, including those used to control for confounding | 9 |
|  |  | (b) Describe any methods used to examine subgroups and interactions |  |
|  |  | (c) Explain how missing data were addressed | 8 |
|  |  | (d) If applicable, describe analytical methods taking account of sampling strategy |  |
|  |  | (e) Describe any sensitivity analyses |  |
| **Results** |  |  |  |
| Participants | 13* | (a) Report numbers of individuals at each stage of study—eg numbers potentially eligible, examined for eligibility, confirmed eligible, included in the study, completing follow-up, and analysed |  |
|  |  | (b) Give reasons for non-participation at each stage |  |
|  |  | (c) Consider use of a flow diagram |  |
| Descriptive data | 14* | (a) Give characteristics of study participants (eg demographic, clinical, social) and information on exposures and potential confounders | 10 |
|  |  | (b) Indicate number of participants with missing data for each variable of interest |  |
| Outcome data | 15* | Report numbers of outcome events or summary measures | 11 |
| Main results | 16 | (a) Give unadjusted estimates and, if applicable, confounder-adjusted estimates and their precision (eg, 95% confidence interval). Make clear which confounders were adjusted for and why they were included | 11-13 |
|  |  | (b) Report category boundaries when continuous variables were categorized | 11-13 |
|  |  | (c) If relevant, consider translating estimates of relative risk into absolute risk for a meaningful time period |  |
| Other analyses | 17 | Report other analyses done—eg analyses of subgroups and interactions, and sensitivity analyses | Repeated measures analysis, Contrast between correlation coefficients |
| **Discussion** |  |  |  |
| Key results | 18 | Summarise key results with reference to study objectives | 14 |
| Limitations | 19 | Discuss limitations of the study, taking into account sources of potential bias or imprecision. Discuss both direction and magnitude of any potential bias | 17-18 |
| Interpretation | 20 | Give a cautious overall interpretation of results considering objectives, limitations, multiplicity of analyses, results from similar studies, and other relevant evidence | 18 |
| Generalisability | 21 | Discuss the generalisability (external validity) of the study results | 17 |
| **Other information** |  |  |  |
| Funding | 22 | Give the source of funding and the role of the funders for the present study and, if applicable, for the original study on which the present article is based | 2 |

\*Give information separately for exposed and unexposed groups.
**Note:** An Explanation and Elaboration article discusses each checklist item and gives methodological background and published examples of transparent reporting. The STROBE checklist is best used in conjunction with this article (freely available on the Web sites of PLoS Medicine at http://www.plosmedicine.org/, Annals of Internal Medicine at http://www.annals.org/, and Epidemiology at http://www.epidem.com/). Information on the STROBE Initiative is available at www.strobe-statement.org.

## Notes

**CONFLICT OF INTEREST** The authors declare that they have no conflict of interest

### Competing Interest Statement

The authors have declared no competing interest.

### Funding Statement

This study did not receive any funding

### Author Declarations

The study was approved by the Clinical Research Ethics Committee of Cantabria (code 2021.102)

